# Immune responses of the third dose of AZD1222 vaccine or BNT162b2 mRNA vaccine after two doses of CoronaVac vaccines against Delta and Omicron variants

**DOI:** 10.1101/2022.10.02.22280572

**Authors:** Suvimol Niyomnaitham, Anan Jongkaewwattana, Atibordee Meesing, Sarunyou Chusri, Sira Nanthapisal, Nattiya Hirankarn, Sarawut Siwamogsatham, Suppachok Kirdlarp, Romanee Chaiwarith, Saranath Lawpoolsri Niyom, Arunee Thitithanyanont, Pokrath Hansasuta, Kanokwan Pornprasit, Sansanee Chaiyaroj, Punnee Pitisuttithum

**Affiliations:** Department of Pharmacology, Faculty of Medicine Siriraj Hospital, Mahidol University, Bangkok, Thailand; National Center for Genetic Engineering and Biotechnology, National Science and Technology Development Agency, Pathumthani, Thailand; Faculty of Medicine, Khon Kaen University, Khon Kaen, Thailand; Clinical research center, Faculty of Medicine, Prince of Songkla University, Songkhla, Thailand; Clinical Research center, Faculty of Medicine, Thammasat University, Pathumthani, Thailand; Department of Microbiology, Faculty of Medicine, Chulalongkorn University, Bangkok, Thailand; Maha Chakri Sirindhorn Clinical Research Center, Faculty of Medicine, Chulalongkorn University, Bangkok, Thailand; Chakri Naruebodindra Medical Institute, Faculty of Medicine Ramathibodi Hospital, Mahidol University, Samut Prakarn, Thailand; Division of Infectious Diseases and Tropical Medicine, Department of Internal Medicine, Faculty of Medicine, Chiang Mai University, Chiang Mai, Thailand; Faculty of Tropical Medicine, Mahidol University, Bangkok, Thailand; Faculty of Science, Mahidol University, Bangkok, Thailand; Division of Virology, Department of Microbiology, Faculty of Medicine, Chulalongkorn University, Bangkok, Thailand; Clinixir Co., Ltd., Bangkok, Thailand; Department of Microbiology, Faculty of Science, Mahidol University, Bangkok, Thailand; Vaccine Trial Centre, Faculty of Tropical Medicine, Mahidol University, Bangkok, Thailand

**Author notes:** These authors contributed equally (Suvimol Niyomnaitham, Anan Jongkaewwattana). **Competing Interest Statement.** All authors report no relevant disclosures. **Funding Statement:** Funding was provided by the Program Management Unit for Competitiveness Enhancement (PMU-C) National research, National Higher Education, Science, Research and Innovation Policy Council, Thailand through Clinixir Ltd. **Role of the Funding Source:** PMU-C funded the trial but had no other involvement in the study. Clinixir provided logistical and monitoring supports. **Author Contributions:** The authors were involved in the study design; in the interpretation of data; in reviewing the manuscript; and in the decision to submit the manuscript for publication. All authors had full access to all of the data in the study and can take responsibility for the integrity of the data and the accuracy of the data analysis. ClinicalTrials.gov Identifier: NCT05049226 The protocol has received approval from Central and each Institutional Ethical Review Boards. All participants had signed consent forms before enrolling into the study.

## Abstract

**Summary:** Half-dose AZD1222 or BNT162b2 boosters maintained immunogenicity and safety, and were non-inferior to full doses. All doses elicited high immunogenicity and best with extended post-CoronaVac primary-series intervals (120-180 days) and high-transmissibility Omicron.

**Methods:** At 60-to-<90, 90-to-<120, or 120-to-180 days (‘intervals’) post-CoronaVac primary-series, participants were randomized to full-dose or half-dose AZD1222 or BNT162b2, and followed up at day-28, -60 and -90. Vaccination-induced immunogenicity to Ancestral, Delta and Omicron BA.1 strains were evaluated by assessing anti-spike (‘anti-S’), anti-nucleocapsid antibodies, pseudovirus neutralization (‘PVNT’), micro-neutralization titers, and T-cells assays. Descriptive statistics and non-inferiority cut-offs were reported as geometric mean concentration (GMC) or titer (GMT) and GMC/GMT ratios comparing baseline to day-28 and day-90 seroresponses, and different intervals post-CoronaVac primary-series. Omicron immunogenicity was only evaluated in full-dose recipients.

**Findings:** No serious or severe vaccine-related safety events occurred. All assays and intervals showed non-inferior immunogenicity between full-doses and half-doses. However, full-dose vaccines and/or longer, 120-to-180-day intervals substantially improved immunogenicity (in GMC measured by anti-S assays or GMT measured by PVNT50; p <0.001). Within platforms and regardless of dose or platform, seroconversions were over 97%, and over 90% for pseudovirus neutralizing antibodies, but similar against the SARS-CoV-2 strains. Immunogenicity waned more quickly with half-doses than full-doses between day 60-to-90 follow-ups, but remained high against Ancestral or Delta strains. Against Omicron, the day-28 immunogenicity increased with longer intervals than shorter intervals for full-dose vaccines.

**Interpretation:** Combining heterologous schedules, fractional dosing, and extended post-second dose intervals, broadens population-level protection and prevents disruptions, especially in resource-limited settings.

**Funding:** Funding was provided by the Program Management Unit for Competitiveness Enhancement (PMU-C) National research, National Higher Education, Science, Research and Innovation Policy Council, Thailand through Clinixir Ltd.

**Research in Context:** *Evidence before this study:*

1. Although nAb titers from CoronaVac primary series waned after 3-4 months, nAb were more increased when boosted at 8 months than at 2 months post-primary series.
2. Six months post-vaccination with a one-fourth dose of primary mRNA-1273, nAb responses were half as robust as full doses, but VE was over 80% of that of full-dose vaccinations.
3. Thai adults boosted with 30μg-BNT162b2 and 15μg-BNT162b2 at 8-12 weeks after two-dose CoronaVac or AZD1222 had high antibodies to the virus receptor-binding domain, nAb titers against all variants, and T-cell responses.
4. Third-dose boosting at a 44–45-week interval significantly increased antibody levels compared to boosting at 15-25-week or 8-12-week intervals.
5. A third dose of CoronaVac administered eight months after the second dose increased antibody levels more than when administered at two months, while antibody responses were two-fold higher with a booster dose of AZD1222 administered at a 12-weeks or longer interval than a 6-weeks or shorter interval.^**Error! Bookmark not defined**^.
6. In the UK, third doses of AZD1222 led to higher antibody levels that correlated with high efficacy and T-cell responses, after a prolonged, dose-stretched interval between vaccine doses, than shorter intervals.
7. Omicron-neutralizing antibodies were detected in only 56% of short-interval vaccine recipients versus all (100%) prolonged-interval vaccine recipients, 69% of whom also demonstrated Omicron-neutralizing antibodies at 4-6 months post-booster.
8. Israeli studies noted a restoration of antibody levels and enhanced immunogenic protection against severe disease when a second booster (fourth dose) was given 4 months or longer after a first booster, with no new safety concerns.

*Added value of this study:* There were no studies designed specifically aimed to analyzed non inferiority between the full dose and half dose of AZD1222 or BNT162b2 boosters after CoronaVac two doses which is important research question when we started the study and the situation of limited vaccine supply, global inequity and high disease burden in the Lower middle-income countries Data on the optimal prime-boost interval is limited, especially data that combines lower (fractional) dosing from resource-limited countries, which is provided by our study.

*Implications of all the available evidence:* We confirm the feasibility of a booster strategy that accounts for the needs of resource-limitations, through the use of fractional dosing, dose-stretching and heterologous schedules, which can broaden population-level protection and prevent vaccination disruptions.

## Introduction

COVID-19 vaccines approved in Thailand include the mRNA-based vaccines, BNT162b2 (‘PF’; Pfizer Inc, New York, United States, US; BioNTech Manufacturing GmbH, Mainz, Germany) and mRNA-1273 (Moderna Inc, Cambridge, US); the inactivated virus vaccine, CoronaVac (Sinovac Biotech, Beijing, China); and the adenovirus vector vaccine, AZD1222 (‘AZ’; Oxford-AstraZeneca, United Kingdom, UK). However, variants of concern like Delta (B.1.617.2) and Omicron, especially BA.1 (B.1.1.529), are driving infection surges amid waning vaccine effectiveness (VE)^1^. Although Thailand has administered over 136 million vaccine doses, Omicron has increased deaths slightly among elderly, bedridden individuals, highlighting a need to sustain vaccinations. Thailand used CoronaVac widely but its immunogenicity and VE are inconsistent or limited.^2^ In China^3^ and Chile^4^, 2-dose CoronaVac (known as ‘primary series’) neutralizing antibody (nAb) titres were lowest against Delta, resulting in breakthrough infections, severe COVID-19 disease and deaths. Although nAb titres from CoronaVac primary-series waned after 3-4 months, nAb were increased more substantially when boosters were administered at 8 months than at 2 months post-primary series^5^.

In 2021, Thailand initiated heterologous regimens with AZD1222, BNT162b2 or mRNA-1273, but was limited by mRNA vaccine shortages. While COVID-19 vaccines are widely available in many Western countries, vaccine inequity and supply constraints remain problematic in many low- and middle-income countries. Thus, different vaccination strategies should be explored, including extended dosing intervals (or dose-stretching), dose reductions or fractional dosing, and vaccine platform combinations. Fractional dosing conserves supplies while increasing coverage without compromising immunogenicity, and can help resource-limited countries extend supplies and reduce mortality.^6,7^ Fractional dosing may also broaden population coverage and facilitate herd immunity, as has been proven in polio. In the COVID-19 pandemic, 6 months post-vaccination with a fractional one-fourth dose of primary mRNA-1273 regimens, nAb responses were half as robust as full doses, but VE was over 80% of that of full-dose vaccinations^8^. During the AZD1222 trials in UK recipients inadvertently primed with half doses and subsequently administered full-dose boosters, VE was 90% (95%CI: 67-97).^9^ Thai adults boosted with 30μg-BNT162b2 and 15μg-BNT162b2 8-12 weeks after two-dose CoronaVac or AZD1222, had high anti-receptor-binding domain (anti-RBD) IgG concentrations^10^, nAb titres against all variants, and T-cell responses^11,12^. Reducing vaccine doses does not necessarily reduce VE, and lower doses could quickly increase the immunity of at-risk populations when vaccine supply is low or during serious disease outbreaks and epidemics. To this end, Moderna has now halved the dose of its mRNA-1273 vaccine from 100-μg to 50-μg, without compromising immunogenicity.^8,13^

Prolonged or different booster intervals may also enhance immunogenicity.^14,15^ In one study, third-dose boosting at 44–45-week intervals significantly increased antibody levels versus boosting at 15-25-week or 8-12-week intervals. Alternatively, SARS-CoV-2 immune responses can also be restored using different booster intervals. A third dose of CoronaVac^16^ administered 8 months after the second dose increased antibody levels more than when administered at 2 months, while antibody responses were 2-fold higher with a booster dose of AZD1222 administered at 12-weeks or longer intervals than 6-weeks or shorter intervals.^15^

In at-risk populations in the United Kingdom, third doses of AZD1222 led to higher antibody levels that correlated with high efficacy and T-cell responses after a prolonged, dose-stretched interval between doses, than at shorter intervals.^14^ In fact, the odds ratios for symptomatic disease following shorter intervals between second and booster (third) doses^17^ was higher than at longer intervals, along with VE estimates of 93.2% (95% CI, 92.8–93.6) versus 95.6% (95% CI, 94.9–96.1), respectively. A Chinese trial of the ZF2001 vaccine^18^ found that it may be feasible to combine these strategies, as ZF2001 neutralizing activity and resilience to all tested variants was higher with extended prime-boost (third dose) intervals, than with shorter intervals. Extended intervals may allow antibodies to mature for longer, thus enhancing this regimen. In fact, Omicron-neutralizing antibodies were detected in only 56% of short-interval vaccine recipients versus all (100%) prolonged-interval vaccine recipients,^19^ 69% of whom also demonstrated Omicron-neutralizing antibodies at 4-6 months post-booster. Although data remains limited on the most optimal prime-boost interval, Israeli studies^20,21^ noted a restoration of antibody levels and enhanced immunogenic protection against severe disease when a second booster (fourth dose) was given 4 months or longer after a first booster, with no new safety concerns.^22^ The WHO now recommends an interval of 4–6 months post-primary series ^23^, especially in the face of Omicron dominance, as well as heterologous or homologous-schedule boosters (third or more).

To better understand the applicability of these strategies, we compared the immune responses and safety of fractional (half) third-doses of heterologous COVID-19 vaccines (AZD1222 or BNT162b2) to full doses after CoronaVac primary series, and when boosting after three different, extended intervals.

## METHODS

### Study design and participants

This prospective, multi-centre, randomized, observer-blinded Phase 2 study enrolled 1,320 healthy adults aged 20 years or older (Figure 1). Eligible participants were either 60-days-to-<90 days, 90-days-to-<120 days, or 120-to-180 days (‘intervals’) post-receipt of CoronaVac primary-series (21-28 days apart); with no history of fever or symptoms within 7 days of enrolment; and were not pregnant. Written informed consents were obtained prior to all study procedures. Exclusion criteria were a history of COVID-19 infection within 3 months of enrolment; contraindication to ChAdOx1 or BNT162b2; and confirmed or suspected immunosuppressive or immunodeficient state. Briefly, participants were divided into two cohorts comprising 660 individuals each and subsequently into three interval-stratified subgroups. Each subgroup was randomly assigned in a 1:1 ratio to either half-dose or full-dose AZD1222 or BNT162b2 vaccine (abbreviated as AZHD, AZFD, PFHD and PFFD). Participants were monitored post-vaccination for immediate adverse events (AEs), and recorded solicited AEs for up to 7 days. All participants completed five post-vaccination follow-up visits at day 28, 60 and 90, and to assess safety at day 7, 28, 60 and 90. Blood was sampled at baseline and days 28, 60 and 90 to evaluate humoral immunity, and at baseline and day 28 from half of participants per subgroup to evaluate T-cell-mediated immunity. Patients were discontinued before study completion due to withdrawal (participant-initiated, for any reason), loss to follow-up, or sponsor-initiated study termination for administrative or other reasons.

**Figure 1.**
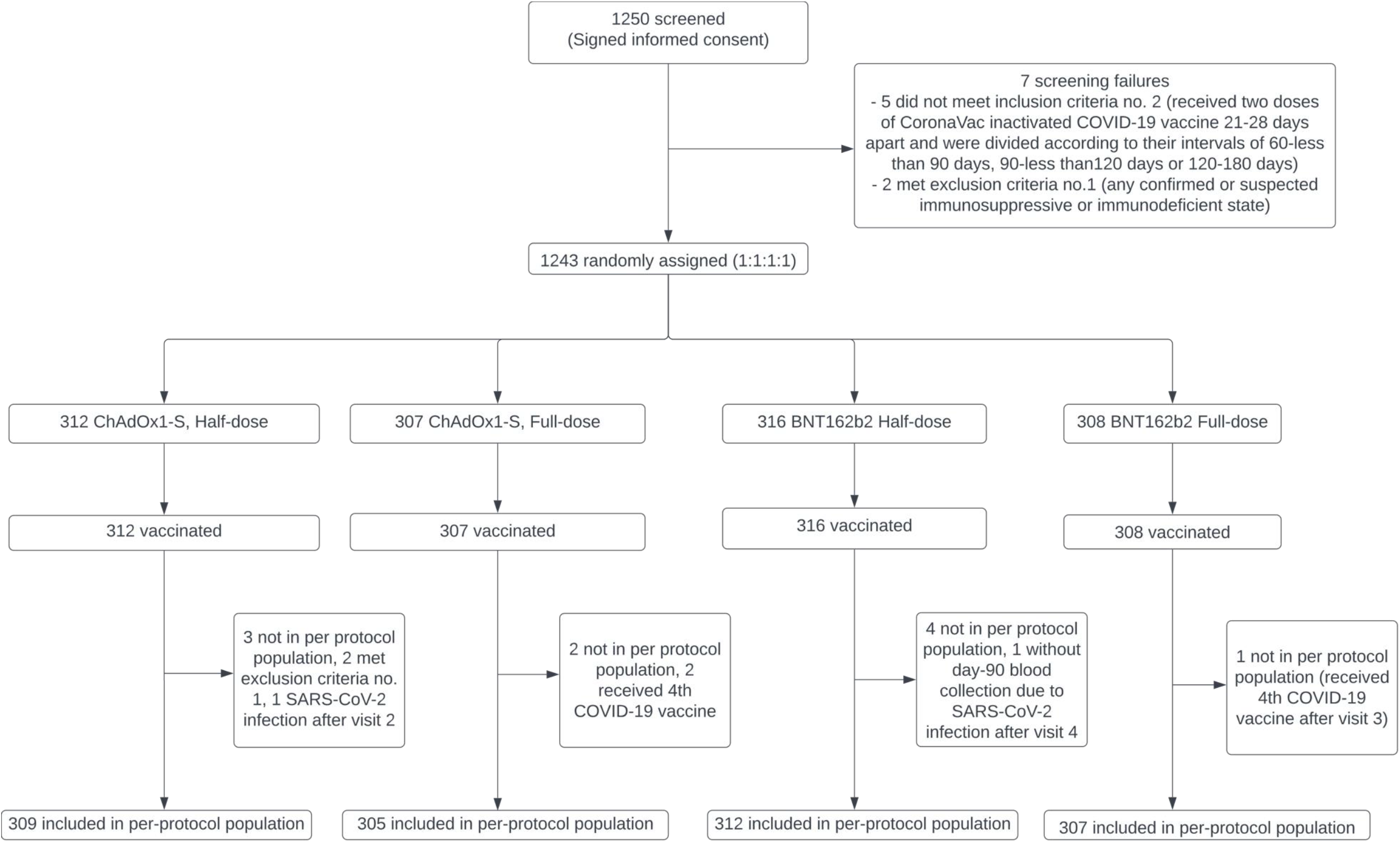
CONSORT diagram depicting trial design and vaccine administration groups.

### Adverse events

Safety analyses included all randomized participants with at least one vaccination dose. All discontinued participants’ diary cards were collected and any AEs and concomitant, ongoing medications at the last visit were recorded. Newly-reported AEs since the last visit were recorded, and AEs were systematically collected at all visits. Diary cards were reviewed at day 7 follow-ups and ongoing solicited AEs were recorded and monitored. AEs of special interest (AESI), medically-assisted AEs (MAAEs), and serious AEs (SAE) were collected through day 90.

### Immunological Analyses

At baseline and days 28, 60 and 90, IgG antibodies to full-length pre-fusion spike protein of SARS-CoV-2 (anti-S RBD IgG) and anti-nucleocapsid (anti-N) proteins levels were quantified using a validated enzyme-linked immunosorbent assay (ELISA; Roche). Anti-S and anti-N RBD IgG results were summarized with 95% exact confidence interval (95%CI), geometric mean concentrations (GMC), geometric mean fold rise (GMFR) from baseline, percentages of participants achieving IgG seroresponse, and reaching the 95%CI for a ≥4-fold increase from baseline. Vaccination-induced nAb inhibition of SARS-CoV-2 seroresponse was quantified by SARS-CoV-2 pseudovirus neutralization assay (PNA), and defined as over 50% and 68% inhibition of Delta and the Ancestral strain, respectively, from baseline, at 28 and 90 days. Samples with >90% inhibition or 100% inhibition were evaluated for 50%-neutralizing titre (NT50) of geometric mean titres (GMT) against Ancestral, Delta or Omicron pseudovirus spike proteins (PVNT). PVNT results were summarized from baseline with 95%CI, GMT, GMFR, and percentage of subjects with ≥4-fold increase in NT50 seroresponse against SARS-CoV-2 pseudovirus from baseline, at 28 and 90 days. SARS-CoV-2 micro-neutralization (microNT) was assayed in pre-screening samples stratified by PVNT against Delta, Ancestral or Omicron. N-proteins were measured by ELISA. SARS-CoV-2 cellular responses (i.e., T-cell-mediated immunity) were quantified using an interferon-gamma release ELISA assay (Euroimmun, Lubeck, Germany).

The primary outcome of immunogenicity was assessed through IgG levels against Anti-S RBD, at baseline, 28, 60 and 90 days after the third-dose/booster given at different intervals among participants with CoronaVac primary series. The immunogenicity, or functional (neutralizing) humoral immune response, elicited by each regimen was also assessed by the PNA at baseline, 28, and 90 days post-third dose. Third-dose vaccination safety and tolerability were evaluated at different intervals among participants with CoronaVac primary-series. The secondary outcomes of functional (neutralizing) humoral immune response at baseline, 28, 90 days was measured by the microNT assay in specimens from participants with ≥4-fold seroconversion, with NT50 GMT assessed after third-dose vaccinations. The exploratory outcome – immunogenicity – was evaluated through S-protein-specific T-cell responses at baseline and 28 days after post-third dose.

### Statistical analyses

Briefly, 110 participants were estimated to be required per arm. Immune response was assessed using two-sided statistical testing with a significance level of 0.05, and a 95%CI. Baseline demographics were summarized for per-protocol (PP) populations using descriptive statistics. Immunogenicity GMFR was computed using estimates of the log difference of the paired samples. Anti-S RBD IgG antibody concentrations were summarized using GMCs at baseline, 28-, 60- and 90-days post-vaccination, GMFR from baseline, and percentage of subjects with IgG seroresponse (all with 95%CI). NT50 nAb titres against SARS-CoV-2 pseudovirus were summarized using GMT at baseline, 28, 90 days post-vaccination, GMFR from baseline, and percentage of subjects with NT50 seroresponse against SARS-CoV-2 pseudovirus at 28- and 90-days post-vaccination (all with 95%CI). Safety was assessed via the Clopper-Pearson method. NT50 nAb titres (seroresponse) against SARS-CoV-2 micro-neutralization titre (microNT) were summarized at baseline, 28- and 90-days post-vaccination using GMT, GMFR from baseline, and percentage of subjects with NT50 seroresponse at 28- and 90-days post-vaccination (all with 95%CI). Non-inferiority of seroresponse rates was calculated between full- and half-dose (AZ and PF) groups. GMC ratios were concluded to be non-inferior when the lower bound of the two-sided 95%CI comparing vaccine groups was >0.67 and the point estimate (PE) was >0.8. The difference in percentage of individuals with ≥4-fold GMFR was concluded to be non-inferior when the lower bound of the two-sided 95%CI for the difference in proportions between vaccine groups was greater than -10%. The difference in percentage of individuals with seroresponse was concluded to be non-inferior when the lower bound of the two-sided 95%CI for the difference in proportions between vaccine groups was greater than -10%. To achieve 80% power to demonstrate non-inferiority it is estimated that 247 subjects per group would be required.

## RESULTS

Between 24th September and 14th October 2021, 1243 of 1250 screened individuals were recruited (Figure 1), randomized (AZHD n=312, AZFD n=307, PFHD n=316, and PFFD n=308; Table 1), administered the planned single-dose of vaccine and included in the safety analysis (SA). Eight participants (0.6%) discontinued (AZHD n=2, AZFD n=3, PFHD n=3, and PFFD n=0), most commonly due to ‘Other: subject inconvenient to come into site’ (n=4). One participant was excluded due to developing COVID-19 (onset day 80). After excluding equal numbers of participants (n=5) in the AZD1222 and BNT162b2 groups, 309 AZHD, 305 AZFD, 312 PFHD, and 307 PFFD participants were included in the per-protocol (PP) set. Exclusions were due to receipt of fourth vaccines (n=6), failed eligibility criteria (n=2), and SARS-CoV-2 infection (n=2). Given the negligible difference (0.8%) in participants in the full and PP analyses, it was reasonable to limit discussions of baseline and immunogenicity to the PP set. The PP population were a mean of 41-years-old and 57% female. Participants were 60-to-<90-days (33%), 90-to-<120 days (34%), or 120-to-180 days (33%) post-final CoronaVac dose.

**Table 1.** Participant demographics of (A) per protocol population, including their (B) medical history and (C) baseline characteristics stratified by vaccine regimen (C), and (D) anti-SARS-CoV-2 nucleocapsid antibody levels at baseline.

| A. Patient Demographics |  |  |  |  |  |
| --- | --- | --- | --- | --- | --- |
|  | ChAdOX1-S |  | BNT162b2 |  | All Subjects (N=1241) |
|  | Half Dose (N=310) | Full Dose (N=307) | Half Dose (N=316) | Full Dose (N=308) |  |
| Age (years) |  |  |  |  |  |
| n (missing) | 310 (0) | 307 (0) | 316 (0) | 308 (0) | 1241 (0) |
| Mean (SD) | 41.7 (9.6) | 41.0 (9.6) | 40.5 (9.8) | 41.9 (9.8) | 41.3 (9.7) |
| Median | 42.0 | 41.0 | 41.0 | 42.0 | 41.0 |
| Age Categories, n (%) |  |  |  |  |  |
| n (missing) | 310 (0) | 307(0) | 316(0) | 308(0) | 1241(0) |
| 20-59 | 303 (97.7) | 301 (98.0) | 315 (99.7) | 304 (98.7) | 1223(98.5) |
| >=60 | 7 (2.3) | 6 (2.0) | 1 (0.3) | 4 (1.3) | 18(1.5) |
| Gender, n (%) |  |  |  |  |  |
| n (missing) | 310 (0) | 307(0) | 316(0) | 308(0) | 1241(0) |
| Male | 116 (37.4) | 139 (45.3) | 133 (42.1) | 141 (45.8) | 529 (42.6) |
| Female | 194 (62.6) | 168 (54.7) | 183 (57.9) | 167 (54.2) | 712 (57.4) |

| B. Medical History |  |  |  |  |  |
| --- | --- | --- | --- | --- | --- |
| System Organ Class<br>Preferred Term | ChAdOX1-S |  | BNT162b2 |  | All Subjects<br>(N=1243) n(%) |
|  | Half Dose<br>(N=312) n(%) | Full Dose<br>(N=307) n(%) | Half Dose<br>(N=316) n(%) | Full Dose<br>(N=308) n(%) |  |
| Metabolism and<br>nutrition disorders | 34(10.9) | 27(8.8) | 32(10.1) | 27(8.8) | 120(9.7) |
| Vascular disorders | 25(8.0) | 27(8.8) | 24(7.6) | 28(9.1) | 104 (8.4) |
| Respiratory, thoracic<br>and mediastinal<br>disorders | 9(2.9) | 12(3.9) | 12(3.8) | 10(3.2) | 43 (3.5) |
| Nervous system<br>disorders | 11(3.5) | 8(2.6) | 9(2.8) | 8(2.6) | 36(2.9) |

| C. Study Vaccine Exposure |  |  |  |  |  |
| --- | --- | --- | --- | --- | --- |
|  | ChAdOX1-S |  | BNT162b2 |  |  |
|  | Half Dose<br>(N=312) | Full Dose<br>(N=307) | Half Dose<br>(N=316) | Full Dose<br>(N=308) | All Subjects (N=1241) |
| Duration of Exposure (days) |  |  |  |  |  |
| n (missing) | 312 (0) | 307 (0) | 316 (0) | 308 (0) |  |
| Mean (SD) | 1.00 (0.00) | 1.00 (0.00) | 1.00 (0.00) | 1.00 (0.00) |  |
| Median | 1.00 | 1.00 | 1.00 | 1.00 |  |
| Number of Injections |  |  |  |  |  |
| n (missing) | 312 (0) | 307 (0) | 316 (0) | 308 (0) |  |
| Mean (SD) | 1.00 (0.00) | 1.00 (0.00) | 1.00 (0.00) | 1.00 (0.00) |  |
| Median | 1.00 | 1.00 | 1.00 | 1.00 |  |
| Cumulative Dose (mL) |  |  |  |  |  |
| n (missing) | 312 (0) | 307 (0) | 316 (0) | 308 (0) |  |
| Mean (SD) | 0.25 (0.00) | 0.50 (0.00) | 0.15 (0.00) | 0.30 (0.00) |  |
| Median | 0.25 | 0.50 | 0.15 | 0.30 |  |
| Interval Duration (days) |  |  |  |  |  |
| n (missing) | 310(0) | 307(0) | 316(0) | 308(0) | 1241(0) |
| 60 to <90 | 102(32.9) | 99(32.2) | 104(32.9) | 103(33.4) | 408(32.9) |
| 90 to <120 | 108(34.8) | 105(34.2) | 111(35.1) | 105(34.1) | 429(34.6) |
| 120 to 180 | 100(32.3) | 103(33.6) | 101(32.0) | 100(32.5) | 404(32.6) |
| D. Baseline Anti-SARS-CoV-2 Nucleocapsid (Anti-N) Antibody |  |  |  |  |  |
| Baseline Anti-N |  |  |  |  |  |
| n (missing) | 46 (264) | 47 (260) | 48 (268) | 44 (264) | 185 (1056) |
| Mean (SD) | 12.03 (40.19) | 12.22 (31.56) | 8.48 (28.50) | 5.41 (16.04) | 9.58 (30.32) |
| Median | 1.8 | 1.5 | 1.6 | 1.3 | 1.6 |

Unsolicited AEs occurred in 38% of participants, including vascular (hypertension and tachycardia), cardiac disorders, and general and administration site conditions (pyrexia). Solicited AE occurred in 84% of participants (Figure 2), including local and systemic reactions (injection site pain, musculoskeletal and connective tissue disorders (fatigue, myalgia, arthralgia), and nervous system disorders (headache). Post-AZHD, three participants experienced unsolicited serious adverse events (SAE) – appendicitis, SARS-CoV-2 infection and haemorrhoids, but all were unrelated to the vaccine and all participants recovered. Nineteen participants (five solicited and 24 unsolicited) experienced medically attended adverse events (MAAE). Two participants had an AESI SARS-CoV-2 infection post-AZHD or post-PFHD and both recovered, but all AESI were unsolicited and unrelated to the vaccine. One PFHD recipient withdrew due to moderate AEs. Neither dose of AZD1222 or BNT2162b2 was directly related to an AE or led to reports of thrombosis with thrombocytopenia syndrome or myocarditis.

**Figure 2.**
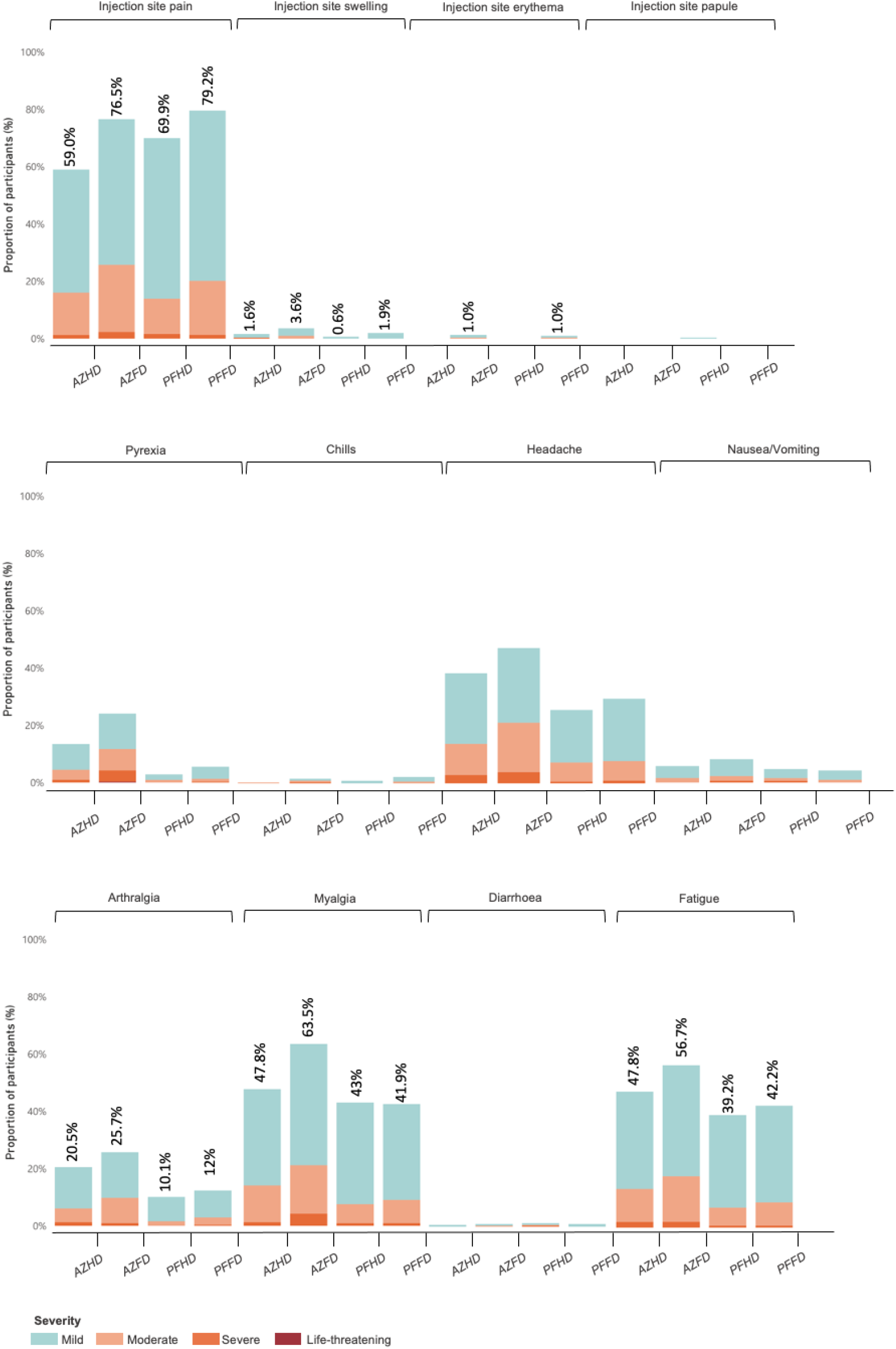
Solicited treatment-emergent adverse events. Abbreviations: AZHD (ChAdOx1-S half-dose) and AZFD (ChAdOx1-S full-dose): AZhalf/AZfull; PFHD (BNT162b2 half-dose) and PFFD (BNT162b2 full-dose): PFhalf/PFfull.

Non-inferiority criteria were met (Figure 3) across all three intervals in comparisons of full-dose to half-dose vaccinations in four immunogenicity assays, and maintained at each interval. To summarise, more than 97% seroconversion rates (≥4-folds GMFR within vaccine types using anti-S IgG) were observed at 28-, 60- and 90-days post-vaccination regardless of dose (Figure 3 and Table 2a). The GMC ratio for anti-S was also non-inferior for AZHD vs AZFD and PFHD vs PFFD (Table 2b).

**Table 2a.** Immunogenicity according to overall anti-spike RBD IgG activity, measured in terms of geometric means concentration (U/mL), geometric mean fold rise and percentage of participants with a more than 4-fold rise in GMFR. Abbreviations: PE, point estimate; CI, confidence interval; AZHD, ChAdOx1-S half-dose; AZFD, ChAdOx1-S full-dose; PFHD, BNT162b2 half-dose; PFFD, BNT162b2 full-dose; GMC, geometric means concentration; GMFR, geometric mean fold rise.

| Analysis Visit | Statistics | AZHD |  | AZFD |  | PFHD |  | PFFD |  |
| --- | --- | --- | --- | --- | --- | --- | --- | --- | --- |
|  |  | PE | 95% CI | PE | 95% CI | PE | 95% CI | PE | 95% CI |
| Baseline | N | 310 |  | 307 |  | 316 |  | 308 |  |
|  | GMC (U/mL) | 48.5 | 43.0, 54.7 | 43.7 | 38.5, 49.6 | 45.7 | 40.3, 51.9 | 44.8 | 39.8, 50.4 |
| Day 28 | N | 309 |  | 307 |  | 315 |  | 308 |  |
|  | GMC (U/mL) | 8237.0 | 7679.3, 8835.2 | 8973.6 | 8328.1, 9669.2 | 14073.9 | 13236.4, 14964.4 | 15920.7 | 15135.7, 16746.4 |
|  | GMFR | 166.9 | 147.8, 188.5 | 205.3 | 181.8, 231.9 | 308.2 | 274.7, 345.7 | 355.5 | 316.0, 400.0 |
|  | ≥4-fold GMFR (%) | 99.4 | 97.7, 99.9 | 99.0 | 97.2, 99.8 | 99.4 | 97.7, 99.9 | 99.7 | 98.2, 100.0 |
| Day 60 | N | 308 |  | 307 |  | 315 |  | 307 |  |
|  | GMC (U/mL) | 5088.3 | 4718.0, 5487.8 | 5496.2 | 5074.3, 5953.1 | 8150.6 | 7584.3, 8759.3 | 9340.6 | 8772.6, 9945.5 |
|  | GMFR | 103.0 | 90.9, 116.7 | 125.7 | 111.0, 142.4 | 177.8 | 157.9, 200.3 | 207.8 | 183.9, 234.8 |
|  | ≥4-fold GMFR (%) | 99.0 | 97.2, 99.8 | 99.0 | 97.2, 99.8 | 99.0 | 97.2, 99.8 | 99.0 | 97.2, 99.8 |
| Day 90 | N | 307 |  | 303 |  | 312 |  | 307 |  |
|  | GMC (U/mL) | 3435.3 | 3166.7, 3726.6 | 3651.3 | 3355.8, 3972.8 | 5097.2 | 4716.2, 5508.9 | 5936.1 | 5540.1, 6360.4 |
|  | GMFR | 69.2 | 60.9, 78.6 | 83.8 | 73.8, 95.3 | 111.5 | 98.4, 126.4 | 132.1 | 116.3, 149.9 |
|  | ≥4-fold GMFR (%) | 98.4 | 96.2, 99.5 | 99.0 | 97.1, 99.8 | 99.0 | 97.2, 99.8 | 99.0 | 97.2, 99.8 |

**Table 2b.** Summary of geometric mean concentration ratios (GMC Ratio) of Anti-SARS-CoV2 Spike-RBD antibody by vaccine group. Overall geometric mean titer ratios of Anti-SARS-CoV2 Spike-RBD antibody was stratified by vaccine group and interval after booster (third) vaccination in the per protocol analysis group. Abbreviations: AZHD, ChAdOx1-S half-dose; AZFD, ChAdOx1-S full-dose; PFHD, BNT162b2 half-dose; PFFD, BNT162b2 full-dose

| Anti SARS-CoV2 Spike-RBD antibody | Interval Duration (Days) | GMC ratio (95%CI) |  |
| --- | --- | --- | --- |
|  |  | AZHD/AZFD | PFHD/PFFD |
| Baseline (D1) | Overall | **0.9 (0.8-1.1)** | **1.0 (0.8-1.2)** |
|  | 60 to <90 | 0.7 (0.6-1.0) | 0.9 (0.7-1.2) |
|  | 90 to < 120 | 1.0 (0.8-1.4) | 1.0 (0.7-1.4) |
|  | 120 to 180 | 1.0 (0.7-1.3) | 1.0 (0.7-1.4) |
| 28 days after the third vaccination | Overall | **0.92 (0.83-1.02)** | **0.88 (0.82-0.96)** |
|  | 60 to <90 | 1.01 (0.85-1.20) | 0.91 (0.79-1.05) |
|  | 90 to < 120 | 0.92 (0.79-1.08) | 0.88 (0.77-1.01) |
|  | 120 to 180 | 0.84 (0.71-1.00) | 0.86 (0.77-0.98) |
| 60 days after the third vaccination | Overall | **0.93 (0.83-1.03)** | **0.87 (0.79-0.96)** |
|  | 60 to <90 | 1.04 (0.87-1.25) | 0.91 (0.78-1.07) |
|  | 90 to < 120 | 0.94 (0.79-1.11) | 0.88 (0.75-1.04) |
|  | 120 to 180 | 0.83 (0.69-0.99) | 0.82 (0.71-0.96) |
| 90 days after the third vaccination | Overall | **0.94 (0.84-1.06)** | **0.86 (0.77-0.95)** |
|  | 60 to <90 | 1.09 (0.89-1.32) | 0.90 (0.75-1.07) |
|  | 90 to < 120 | 0.95 (0.78-1.15) | 0.87 (0.73-1.03) |
|  | 120 to 180 | 0.82 (0.68-1.00) | 0.81 (0.68 - 0.96) |

**Figure 3.**
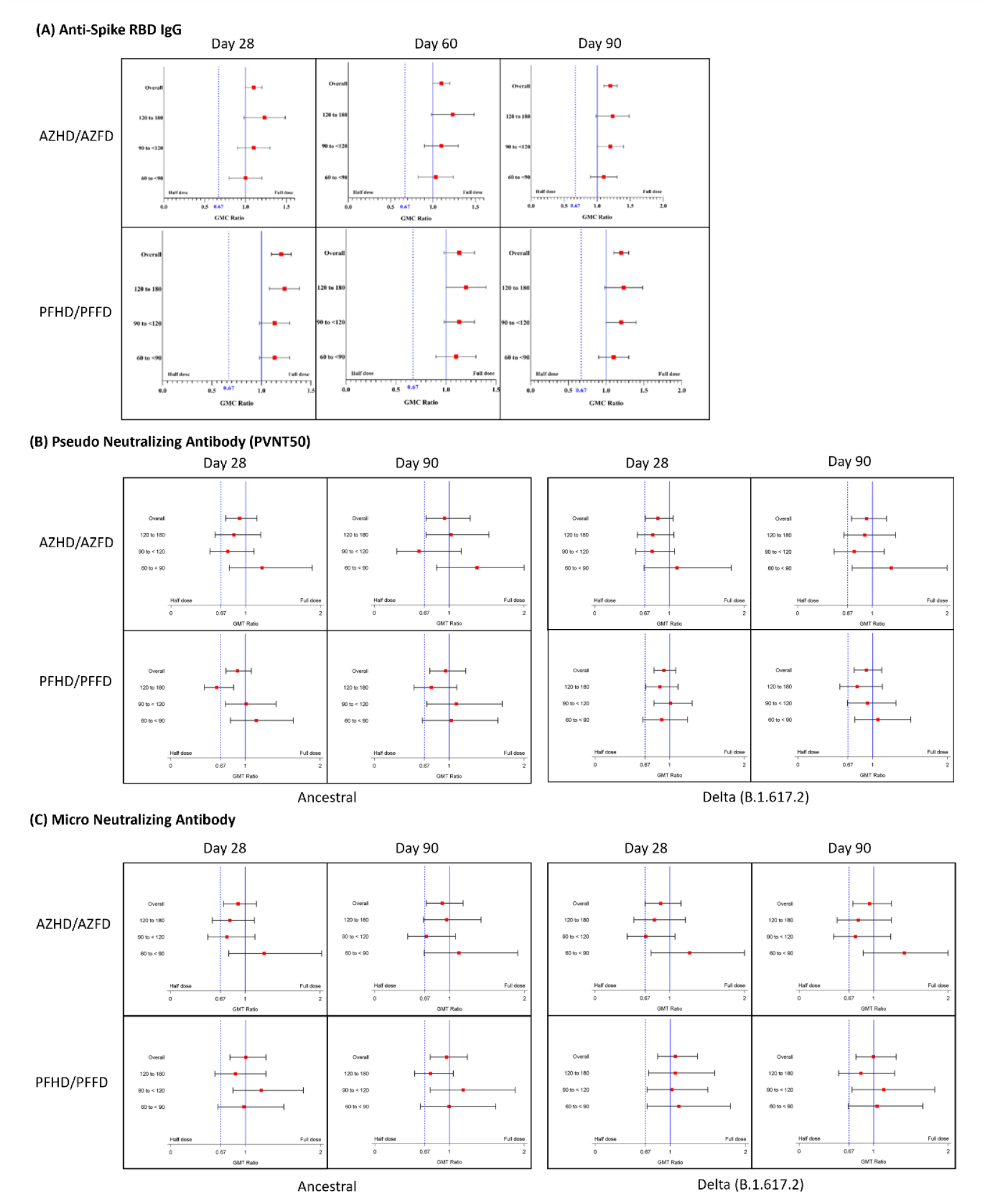
Non-inferiority comparisons for all immunogenicity assays by vaccine group. All comparisons were between either AZHD and AZFD; or between PFHD and PFFD and stratified by interval after booster (third) vaccination in the per protocol analysis group for (A) anti-SARS-Cov-2 Spike RBD antibody (shown as GMC Ratio), (B) pseudovirus neutralization antibody titre (shown as GMT Ratio) against Ancestral SARS-CoV-2 (left) or Delta (B1.617.2) variant (right), and (C) micro-neutralization (shown as GMT Ratio) against Ancestral SARS-CoV-2 (left) or Delta (B.1.617.2) (right). Abbreviations: AZHD, ChAdOx1-S half-dose; AZFD, ChAdOx1-S full-dose; PFHD, BNT162b2 half-dose; PFFD, BNT162b2 full-dose; GMC Ratio, geometric mean concentration ratio; GMT Ratio, geometric mean titre ratio; RBD, receptor-binding domain.

Pseudovirus neutralizing antibodies demonstrated more than 90% seroconversion rates (≥4-fold rise). Moreover, the responses across dose groups were of similar magnitudes between the two vaccine platforms, at 28 days post-vaccination, and against the Ancestral, Delta and Omicron strains (Table 3a, Figure 4). However, GMT and GMFR declined against Delta and Omicron as detailed below (Table 3a). Regardless, non-inferiority in terms of nAb was met for AZHD versus AZFD and for PFHD versus PFFD (Table 3b). Importantly, a longer interval between CoronaVac primary-series and the booster (third) dose (‘prime-boost’) improved immunogenicity. Boosting at 120-to-180-days substantially improved GMC as measured by anti-S IgG and GMT as measured by PVNT50, especially with the full-doses of both platforms against each strain (p <0.001) (Table 2a-2b and Figure 4A).

**Table 3a.** Immunogenicity according to Pseudovirus Neutralizing Antibody Titer (PVNT50) against different variant of concerns, measured in terms of geometric means titer (GMT), geometric mean fold rise (GMFR) and percentage of participants with a more than 4-fold rise in GMFR.

| Analysis Visit | Statistics | AZHD |  | AZFD |  | PFHD |  | PFFD |  |
| --- | --- | --- | --- | --- | --- | --- | --- | --- | --- |
|  |  | PE | 95% CI | PE | 95% CI | PE | 95% CI | PE | 95% CI |
| Ancestral SARS-CoV-2 |  |  |  |  |  |  |  |  |  |
| Baseline | N | 100 |  | 97 |  | 100 |  | 99 |  |
|  | GMT | 24.2 | 21.5-27.2 | 24.6 | 21.5-28.2 | 27.4 | 22.8-33.1 | 25.8 | 22.9-29.0 |
| Day 28 | N | 100 |  | 97 |  | 100 |  | 99 |  |
|  | GMT | 653.2 | 556.9-766.3 | 712.2 | 607.0-835.6 | 1219.3 | 1048.7-1417.6 | 1362.7 | 1207.6-1537.9 |
|  | GMFR | 27.0 | 22.5-32.4 | 28.9 | 24.2-34.7 | 44.4 | 36.5-54.1 | 52.9 | 45.0-62.1 |
|  | ≥4-fold GMFR (%) | 97.1 | 94.5-98.7 | 97.7 | 95.4-99.1 | 100.0 | 98.8-100.0 | 100.0 | 98.8-100.0 |
| Day 90 | N | 100 |  | 97 |  | 98 |  | 99 |  |
|  | GMT | 407.0 | 335.6-493.6 | 406.1 | 333.9-493.9 | 539.6 | 438.9-663.4 | 575.3 | 494.1-669.8 |
|  | GMFR | 16.8 | 13.7-20.7 | 16.5 | 13.2-20.7 | 20.4 | 16.3-25.5 | 22.3 | 18.6-26.8 |
|  | ≥4-fold GMFR (%) | 91.9 | 88.2-94.7 | 94.7 | 91.6-97.0 | 98.1 | 95.9-99.3 | 98.7 | 96.7-99.6 |
| Delta (B.1.617.2) |  |  |  |  |  |  |  |  |  |
| Baseline | N | 190 |  | 187 |  | 213 |  | 202 |  |
|  | GMT | 22.1 | 20.5-23.7 | 23.8 | 21.6-26.3 | 22.8 | 20.9-24.9 | 21.7 | 20.7-22.9 |
| Day 28 | N | 190 |  | 187 |  | 213 |  | 202 |  |
|  | GMT | 468.3 | 409.8-535.2 | 530.6 | 470.4-598.5 | 801.5 | 715.4-897.8 | 856.1 | 777.8-942.1 |
|  | GMFR | 21.2 | 18.3-24.6 | 22.3 | 19.3-25.8 | 35.1 | 30.8-40.0 | 39.4 | 35.4-43.8 |
|  | ≥4-fold GMFR (%) | 90.6 | 86.8-93.6 | 94.1 | 90.9-96.5 | 98.1 | 95.9-99.3 | 99.4 | 97.7-99.9 |
| Day 90 | N | 190 |  | 186 |  | 210 |  | 202 |  |
|  | GMT | 175.1 | 152.6-200.9 | 184.1 | 160.9-210.8 | 221.9 | 194.4-253.3 | 225.4 | 200.9-252.9 |
|  | GMFR | 7.9 | 6.9-9.2 | 7.7 | 6.6-9.0 | 9.8 | 8.5-11.2 | 10.4 | 9.2-11.7 |
|  | ≥4-fold GMFR (%) | 77.5 | 72.4-86.1 | 82.2 | 77.4-86.3 | 86.9 | 82.6-90.4 | 91.2 | 87.5-94.1 |
| Omicron (B.1.1.529) |  |  |  |  |  |  |  |  |  |
| Baseline | N |  |  | 238 |  |  |  | 238 |  |
|  | GMT |  |  | 1.6 | 1.4-1.8 |  |  | 1.5 | 1.3-1.8 |
| Day 28 | N |  |  | 238 |  |  |  | 238 |  |
|  | GMT |  |  | 118.9 | 97.3-145.2 |  |  | 255.9 | 222.9-293.7 |
|  | GMFR |  |  | 76.1 | 59.4-97.6 |  |  | 167.1 | 137.7-202.7 |
|  | ≥4-fold GMFR (%) |  |  | 90.3 | 85.9-93.8 |  |  | 97.1 | 94.0-98.8 |

**Table 3b.** Summary of geometric mean titer ratios (GMT Ratio) of Pseudovirus Neutralizing Antibody Titers (PVNT50) by vaccine group. Overall geometric mean titer ratios of PVNT50 was stratified by vaccine group and interval after booster (third) vaccination in the per protocol analysis group. Abbreviations: AZHD, ChAdOx1-S half-dose; AZFD, ChAdOx1-S full-dose; PFHD, BNT162b2 half-dose; PFFD, BNT162b2 full-dose

| Timepoint of PVNT50 Measurement Against SARS-CoV-2 | Interval Duration (Days) | GMT Ratio (95% CI) |  |  |  |
| --- | --- | --- | --- | --- | --- |
|  |  | Ancestral SARS-CoV-2 |  | Delta (B.1.617.2) SARS-CoV-2 |  |
|  |  | AZHD/AZFD | PFHD/PFFD | AZHD/AZFD | PFHD/PFFD |
| Baseline (D1) | Overall | **0.73 (0.45-1.18)** | **1.25 (0.74-2.10)** | **0.79 (0.55-1.13)** | **1.04 (0.75-1.45)** |
|  | 60 to <90 | 1.12 (0.46-2.69) | 2.88 (1.12-7.36) | 1.55 (0.81-2.98) | 1.40 (0.73-2.69) |
|  | 90 to < 120 | 0.33 (0.15-0.71) | 1.41 (0.58-3.46) | 0.54 (0.29-1.02) | 0.76 (0.44-1.34) |
|  | 120 to 180 | 1.05 (0.44-2.49) | 0.47 (0.20-1.10) | 0.69 (0.39-1.24) | 1.11 (0.66-1.88) |
| 28 days after the third vaccination | Overall | **0.92 (0.73-1.15)** | **0.89 (0.74-1.08)** | **0.88 (0.74-1.06)** | **0.94 (0.81-1.09)** |
|  | 60 to <90 | 1.22 (0.78-1.89) | 1.15 (0.8-1.64) | 1.16 (0.80-1.68) | 0.94 (0.71-1.23) |
|  | 90 to < 120 | 0.76 (0.52-1.11) | 1.01 (0.73-1.41) | 0.80 (0.59-1.07) | 1.01 (0.79-1.30) |
|  | 120 to 180 | 0.84 (0.59-1.20) | 0.62 (0.45-0.84) | 0.81 (0.62-1.06) | 0.87 (0.68-1.11) |
| 90 days after the third vaccination | Overall | **1.00 (0.76-1.32)** | **0.94 (0.73-1.21)** | **0.95 (0.78, 1.15)** | **0.98 (0.83, 1.17)** |
|  | 60 to <90 | 1.40 (0.84-2.34) | 1.01 (0.62-1.64) | 1.25 (0.86-1.82) | 1.10 (0.80-1.51) |
|  | 90 to < 120 | 0.71 (0.43-1.17) | 1.07 (0.68-1.70) | 0.87 (0.61-1.25) | 1.01 (0.75-1.35) |
|  | 120 to 180 | 1.02 (0.69-1.53) | 0.76 (0.53-1.10) | 0.86 (0.65-1.13) | 0.88 (0.66-1.19) |

**Figure 4.**
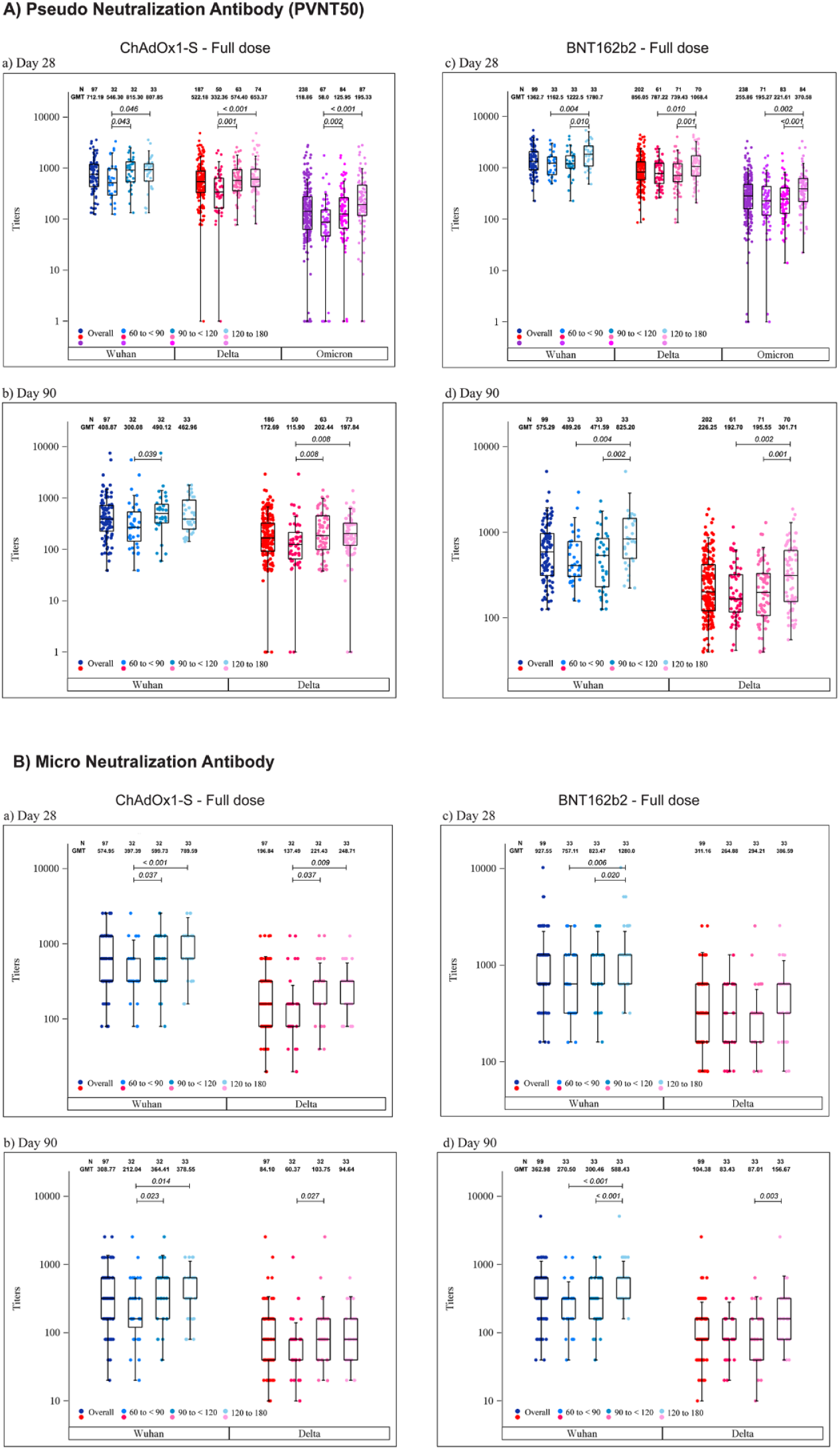
Vaccine-induced immune responses. Geometric mean titres (GMT) of (A) pseudo and (B) micro-neutralization antibody assays are stratified by variant after participant completed primary CoronaVac vaccinations and before administration of boosters.

The GMFR (%) for AZHD was 166.9, 103 and 69.2 at days 28, 60 and 90, respectively, versus 205.3, 125.7 and 83.8 for AZFD at the same respective visits. The GMFR for PFHD was 308.2, 177.8 and 111.5 at days 28, 60 and 90, respectively, versus 355.5, 207.8 and 132.1 for PZFD at the same respective visits. Anti-S RBD IgG GMC PE peaked at day 28 for all doses and vaccine platforms (data not shown; AZHD PE: 8237.0, 95%CI: 7679.3-8835.2; AZFD PE: 8973.6, 95%CI: 8328.1-9669.2; PFHD PE: 14073.9, 95%CI: 13236.4-14964.4; and PFFD PE: 15920.7, 95%CI: 15135.7-16746.4). GMC PE was still high and comparable at day 90 between AZHD and AZFD (3435.3, 95%CI: 3166.7-3726.6, and 3651.3, 95%CI: 3355.8-3972.8), and between PFHD and PFFD (5097.2, 95%CI: 4716.2-5508.9, and 5936.1; 95%CI: 5540.1-6360.4).

At day 28 and 90, the baseline PVNT50 GMT against SARS-CoV-2 Delta and Ancestral strains (Table 3a) was similar between vaccines. However, at day 90, AZHD and AZFD had similar GMT against the Ancestral strain. Regardless of strain, PVNT50 GMT peaked at day 28. At days 28 and 90, AZHD PVNT50 GMT against Delta was non-inferior to AZFD, while PFHD was non-inferior to PFFD (Table 3b). Similarly, AZHD PVNT50 GMT against the Ancestral strain was non-inferior to AZFD, while PFHD was non-inferior to PFFD. Against Delta, the PVNT50 GMT PE for AZHD was 468.3 (95%CI: 409.8-535.2) and 175.1 (95%CI: 152.6-200.9), versus 530.6 (95%CI: 470.4-598.5) and 184.1 (95%CI: 160.8-210.7) for AZFD. The PVNT50 GMT for PFHD was 801.5 (95%CI: 715.4-897.8) and 221.9 (95%CI: 194.4-253.3), versus 856.1 (95%CI: 777.8-942.1) and 225.4 (95%CI: 200.9-252.9) for PFFD. Against the Ancestral strain, PVNT50 GMT for AZHD was 653.2 (95%CI: 556.88-766.28) and 407.0 (95%CI: 335.6-493.6) at days 28 and 90 respectively, versus 712.2 (95%CI: 607.0-835.6) and 406.1 (95%CI: 333.9-493.9) for AZFD. The PVNT50 GMT for PFHD was 1219.3 (95%CI: 1048.7-1417.6) and 539.6 (95%CI: 438.97-663.4), versus 1362.7 (95%CI: 1207.68-1537.9) and 575.3 (95%CI: 494.1-669.8) for PFFD. Strain-specific PVNT50 GMT ratios (Table 3b, Figure 4A) also showed an improved immunogenicity with longer prime-boost intervals (p<0.001). Against Delta, the AZHD/AZFD GMT ratio was high and increased at days 28 and 90, but the PFHD/PFFD GMT ratio, though high, was relatively unchanged between day 28 [0.9 (0.8-1.1)] and 90 [1.0 (0.8-1.2)]. Against the Ancestral strain, the GMT ratios for both AZHD/AZFD and PFHD/PFFD were high and increased at days 28 and 90. When PVNT50 was stratified by boosting interval (Figure 4 A, a-d), greater differences were seen between vaccine types at the 60-to-<90-day interval, with all comparisons inferior for Delta. At the 90-to-<120-day interval all comparisons were non-inferior, whereas at the 120-to-180-days interval, most comparisons were non-inferior. Overall, the Ancestral strain prompted higher seroresponse rates than the Delta strain. Also, seroresponses against Delta were non-inferior at day 28 between dose levels of either vaccine, but seroresponses were only non-inferior between PFHD and PFFD at day 90. For the Ancestral strain, non-inferiority was observed at all time points and within dose levels for each vaccine.

Full-dose vaccine microNT and PVNT50 were consistent across post-baseline visits for Delta and the Ancestral strain, and non-inferiority was observed at all intervals when stratified by variant (Figure 4A and 4B a-d). At day 28, many participants also had ≥4-fold increase in microNT titres to both Delta and the Ancestral strain, with all doses and vaccines. The microNT NT50 against both Delta and the Ancestral strains peaked at day 28 versus day 90 (Table 4a, 4b). Against Delta, the microNT NT50 GMT PE for AZHD was 172.7 (95%CI: 142.3-209.5) versus 196.8 (95%CI: 163.2-237.4) for AZFD at day 28 (p=0.336), and 79.5 (95%CI: 65.6-96.2) versus 84.1 (95%CI: 69.0-102.5), respectively, at day 90 (p=0.681). Similarly, the microNT NT50 GMT for PFHD was 331.3 (95%CI: 274.5-399.8) versus 311.2 (95%CI: 265.7-364.4) for PFFD at day 28 (p=0.613), and 103.9 (95%CI: 84.9-127.2) versus 104.4 (95%CI: 87.5-124.5), respectively, at day 90 (p=0.974). Similar trends were observed with the Ancestral strain: the microNT NT50 GMT for AZHD was 519.8 (95%CI: 435.7-620.2) versus 575.0 (95%CI: 487.3-678.4) for AZFD (p=0.410) at day 28, and 278.6 (95%CI: 231.0-336.0) versus 308.8 (95%CI: 254.4-374.8), respectively, at day 90 (p=0.450). Also, the microNT NT50 GMT for PFHD was 930.5 (95%CI: 780.1-1110.1) versus 927.6 (95%CI: 793.7-1084.0) for PFFD at day 28 (p=0.978), and 348.35 (95%CI: 285.5-4254.0) versus 363.0 (95%CI: 306.5-429.9), respectively, at day 90 (p=0.755). All intervals between doses of each vaccine were also non-inferior between Delta and the Ancestral strain.

**Table 4a.** Immunogenicity according to Micro Neutralizing Antibody Titer (MicroNT) against different variant of concerns, measured in terms of geometric means titer (GMT), geometric mean fold rise (GMFR) and percentage of participants with a more than 4-fold rise in GMFR.

| Analysis Visit | Statistics | AZHD |  | AZFD |  | PFHD |  | PFFD |  |
| --- | --- | --- | --- | --- | --- | --- | --- | --- | --- |
|  |  | PE | 95% CI | PE | 95% CI | PE | 95% CI | PE | 95% CI |
| Ancestral SARS-CoV-2 |  |  |  |  |  |  |  |  |  |
| Baseline | N | 100 |  | 97 |  | 100 |  | 99 |  |
|  | GMT | 23.5 | 19.9-27.7 | 22.1 | 18.9-25.9 | 22.2 | 18.6-26.5 | 21.9 | 18.4-26.2 |
| Day 28 | N | 100 |  | 97 |  | 100 |  | 99 |  |
|  | GMT | 519.8 | 435.7-620.2 | 575.0 | 487.3-678.4 | 930.5 | 780.1-1110.0 | 927.6 | 793.7-1084.0 |
|  | GMFR | 22.2 | 17.9-27.4 | 26.0 | 21.3-31.8 | 41.9 | 33.9-51.8 | 42.3 | 33.9-52.9 |
|  | ≥4-fold GMFR (%) | 98.0 | 93.0-99.8 | 100.00 | 96.3-100.0 | 99.0 | 94.6-100.0 | 99.0 | 94.5-100.0 |
| Day 90 | N | 100 |  | 97 |  | 98 |  | 99 |  |
|  | GMT | 278.6 | 231.0-336.0 | 308.8 | 254.4-374.8 | 348.4 | 285.5-425.0 | 363.0 | 306.5-429.9 |
|  | GMFR | 11.9 | 9.4-15.0 | 14.0 | 11.1-17.7 | 15.7 | 12.4-19.8 | 16.6 | 13.0-21.1 |
|  | ≥4-fold GMFR (%) | 93.0 | 86.1-97.1 | 92.8 | 85.7-97.0 | 91.8 | 84.5-96.4 | 94.9 | 88.6-98.3 |
| Delta (B.1.617.2) |  |  |  |  |  |  |  |  |  |
| Baseline | N | 100 |  | 97 |  | 100 |  | 99 |  |
|  | GMT | 12.1 | 11.0-13.4 | 13.0 | 11.5-14.8 | 12.9 | 11.4-14.7 | 11.7 | 10.6-12.8 |
| Day 28 | N | 100 |  | 97 |  | 100 |  | 99 |  |
|  | GMT | 172.7 | 142.3-209.5 | 196.8 | 163.2-237.4 | 331.3 | 274.5-399.8 | 311.2 | 265.7-364.4 |
|  | GMFR | 14.2 | 11.7-17.3 | 15.1 | 12.2-18.7 | 25.6 | 20.9-31.5 | 26.7 | 22.1-32.2 |
|  | ≥4-fold GMFR (%) | 95.0 | 88.7-98.4 | 92.8 | 85.7-97.1 | 96.0 | 90.1-98.9 | 100.0 | 96.3-100.0 |
| Day 90 | N | 100 |  | 97 |  | 98 |  | 99 |  |
|  | GMT | 79.5 | 65.6-96.2 | 84.1 | 69.0-102.5 | 103.9 | 84.9-127.2 | 104.4 | 87.5-124.5 |
|  | GMFR | 6.5 | 5.4-8.0 | 6.5 | 5.1-8.1 | 8.0 | 6.5-9.9 | 9.0 | 7.4-10.9 |
|  | ≥4-fold GMFR (%) | 81.0 | 71.9-88.2 | 79.4 | 70.0-86.9 | 81.6 | 72.5-88.7 | 88.9 | 81.0-94.3 |

**Table 4b.** Summary of geometric mean titer ratios (GMT Ratio) of Micro Neutralization Antibody Titers (MicroNT) by vaccine group. Overall geometric mean titer ratios of MicroNT was stratified by vaccine group and interval after booster (third) vaccination in the per protocol analysis group. Abbreviations: AZHD, ChAdOx1-S half-dose; AZFD, ChAdOx1-S full-dose; PFHD, BNT162b2 half-dose; PFFD, BNT162b2 full-dose

| Timepoint of MicroNT Measurement Against SARS-CoV-2 | Interval Duration (Days) | GMT Ratio (95% CI) |  |  |  |
| --- | --- | --- | --- | --- | --- |
|  |  | Ancestral SARS-CoV-2 |  | Delta (B.1.617.2) SARS-CoV-2 |  |
|  |  | AZHD/AZFD | PFHD/PFFD | AZHD/AZFD | PFHD/PFFD |
| Baseline (D1) | Overall | **1.06 (0.84-1.33)** | **1.01 (0.79-1.30)** | **0.93 (0.79-1.10)** | **1.11 (0.95-1.29)** |
|  | 60 to <90 | 1.20 (0.79-1.81) | 1.17 (0.76-1.79) | 0.95 (0.67-1.34) | 1.22 (0.92-1.62) |
|  | 90 to < 120 | 0.90 (0.60-1.34) | 0.92 (0.54-1.55) | 0.93 (0.72-1.20) | 0.96 (0.68-1.35) |
|  | 120 to 180 | 1.11 (0.77-1.60) | 0.96 (0.71-1.29) | 0.92 (0.73-1.16) | 1.16 (0.99-1.36) |
| 28 days after the third vaccination | Overall | **0.90 (0.71-1.15)** | **1.00 (0.79-1.27)** | **0.88 (0.67-1.15)** | **1.06 (0.83-1.36)** |
|  | 60 to <90 | 1.25 (0.78-2.02) | 0.97 (0.63-1.51) | 1.27 (0.75-2.12) | 1.11 (0.69-1.80) |
|  | 90 to < 120 | 0.75 (0.50-1.13) | 1.21 (0.83-1.77) | 0.68 (0.43-1.07) | 1.02 (0.69-1.50) |
|  | 120 to 180 | 0.79 (0.56-1.12) | 0.86 (0.59-1.27) | 0.79 (0.52-1.21) | 1.07 (0.71-1.59) |
| 90 days after the third vaccination | Overall | **0.90 (0.69-1.18)** | **0.96 (0.74-1.24)** | **0.94 (0.72-1.24)** | **1.00 (0.76-1.30)** |
|  | 60 to <90 | 1.12 (0.66-1.91) | 0.99 (0.61-1.62) | 1.41 (0.86-2.31) | 1.05 (0.66-1.66) |
|  | 90 to < 120 | 0.69 (0.44-1.08) | 1.18 (0.74-1.88) | 0.76 (0.46-1.23) | 1.13 (0.71-1.82) |
|  | 120 to 180 | 0.96 (0.65-1.42) | 0.75 (0.53-1.05) | 0.79 (0.51-1.24) | 0.83 (0.53-1.28) |

Day-28 T-cell levels (Table 5) were higher with BNT162b2 than AZD1222 (p<0.01), and higher with full-dose BNT162b2 than half-dose BNT162b2 (p=0.022), but similar regardless of AZD1222 dose (p=0.642) or interval duration. Thus, full-dose AZD1222 was non-inferior to half-dose AZD1222, and full-dose BNT162b2 was non-inferior to half-dose BNT162b2.

**Table 5.** SARS-CoV-2 Reactive T-cell Concentration IFN-gamma Assay. Results are presented in units of mIU/mL. Q1, 1^st^ Quartile, Q3, 3^rd^ Quartile

| Analysis Visit | AZHD<br>N=222 |  | AZFD<br>N=231 |  | PFHD<br>N=223 |  | PFFD<br>N=224 |  |
| --- | --- | --- | --- | --- | --- | --- | --- | --- |
|  | Median | Q1-Q3 | Median | Q1-Q3 | Median | Q1-Q3 | Median | Q1-Q3 |
| Baseline | 257.5 | 111.1-623.5 | 228.0 | 110.4-486.0 | 238.1 | 98.7-553.1 | 250.6 | 114.7-572.7 |
| Day 28 | 1722.4 | 838.3-3245.2 | 1620.0 | 968.6-3305.1 | 2866.9 | 1391.6-6398.1 | 3487.7 | 1853.1-6835.7 |

Against Omicron, the day-28 PVNT50 GMT increased with longer booster intervals (Figure 4A) for AZFD and PFFD. A 60-to-<90-day interval increased the PVNT50 GMT PE from 1.4 (95%CI: 1.1-1.8) with AZFD and 1.6 (95%CI: 1.3-2.0) with PFFD to 58.00 (95%CI: 37.7-89.3) and 195.3 (95%CI: 140.4-271.6), respectively. A 90-to-<120-day interval increased the PVNT50 GMT from 1.6 (95%CI: 1.3273-2.1) with AZFD and 1.46 (95%CI: 1.2-1.8) with PFFD to 1260 (95%CI: 93.9-169.0) and 221.6 (95%CI: 184.0-2676.0), respectively. The 120-to-180-day interval increased the PVNT50 GMT from 1.6 (95%CI: 1.3-2.18) with AZFD and 1.5 (95%CI: 1.2-1.9) with PFFD to 195.3 (95%CI: 145.0-263.0) and 370.6 (95%CI: 307.2-447.1), respectively.

## DISCUSSION

This randomized, double-blind study compared the safety and immunogenicity induced by boosting with an additional (third) half-dose to full-dose AZD1222, and half-dose to full-dose BNT162b2, and between three, extended post-primary vaccination intervals. Comparisons were not conducted between vaccine platforms (AZD1222 versus BNT162b2). Instead, our intra-platform comparisons differentiated between dose, boosting intervals and virus variants, with sufficient statistical power to detect non-inferiority between different doses of the same vaccine. We did so in order to understand if and how to maintain high levels of immunogenicity when vaccine doses were limited.

Although our safety assessments were limited by small cohort sizes, all vaccine doses were found to be very safe, with no vaccine-related or life-threatening AE occurring at any dose. Thrombosis with thrombocytopenia syndrome (TTS), though rare, has been documented with AZD1222.^24^ However, no TTS occurred in our study with either AZHD or AZFD. Moreover, no previously-reported BNT162b2 AESI or SAE occurred in our study.^25^

Importantly, half-dose AZD1222 or BNT162b2 were immunologically non-inferior to the corresponding full-dose in anti-spike IgG assays, PNA and microNT assays of humoral immune responses and S-protein-specific IFN-γ assays for T-cell response. The non-inferiority between doses of any vaccine type persisted at all intervals, even 90-120 days after primary-series CoronaVac. While seroconversion was high at all doses, it declined slightly with longer intervals in half-dose recipients, and more slowly in full-dose recipients. Specifically, at day 90, the half-dose immune response waned more than the full-dose immune response; nevertheless, the half-dose immune response at day 28 remained robust and non-inferior. Reassuringly, a high and comparable proportion of participants seroconverted after the different intervals, with increased seroconversion and T-cell induction rates at longer intervals, even at 120 days post-primary series. Also, our full-dose vaccination induced similar immune responses to another Thai study that had a smaller cohort.^26^ A vaccination strategy which allows lower doses can potentially mitigate supply constraints, especially in resource-limited settings facing an urgent need to ensure high vaccination coverage. We showed, in a large, adult population, that half-dose AZ1222 boosting was non-inferior to full-dose AZ1222 boosting, and half-dose BNT162b2 boosting was non-inferior to full-dose BNT162b2 boosting, in terms of eliciting high immunogenicity. This was especially the case with longer intervals between prime-boost schedules, and with the high transmissibility of Omicron. Half-dose vaccinations were not evaluated for Omicron since it is associated with immune escape and must be overcome by full doses.^27^ Notably, virus neutralization declined gradually from the Ancestral strain to the Delta variant, and more markedly from these strains to Omicron. Regardless, neutralization remained high even with extended intervals between primary and booster doses and as recently shown, nAb titres may correlate with protection against infection^28^. Halving doses did not compromise immune responses or safety, and could be an important strategy for ensuring population coverage, especially when boosting. Now, a main vaccine provider, Moderna Inc., is manufacturing 50-μg doses of mRNA-1273, and will produce bivalent boosters with 25 μg of each antigen.^29^

Low or inconsistent vaccine supplies can be circumvented with heterologous regimens. mRNA-1273, BNT162b2 and AZD1222 vaccine combinations are safe, well-tolerated and comparably or even more immunogenic than homologous regimens. A strategy that combines fractional dosing, dose-stretching and heterologous schedules, can prevent vaccination campaign disruptions even if vaccine logistics or safety are problematic. Future investigations should include age-based stratifications and comparative durations of humoral immunity between fractional and full doses. However, with the current spread of Omicron globally, provision of full-dose AZ or PF boosters is consistent with recent effectiveness data.^30^

## CONCLUSION

We found similar immune responses after day 28 when boosting at any of the three, progressively longer intervals post-primary series, and that a third dose given at a longer interval was most optimal. Halving doses does not compromise immunogenicity or safety, but can broaden vaccine coverage when vaccine supply is a problem.

## Data Availability

All data produced in the present study are available upon reasonable request to the authors.

## Acknowledgment

The authors wish to thank the staff of all seven medical schools, Clinixir, Biotec, MHSRI and Faculty of Science, Mahidol University and Shawna Tan of Medical Writers Asia for assisting in drafting manuscript, Professor Kanta Subbarao, Dr.Jean-Louis Excler and Assoc. Prof. Jaranit Kaewkungwal who have served as Data and Safety Monitoring Board.

## Notes

### Competing Interest Statement

The authors have declared no competing interest.

### Clinical Trial

NCT05049226

### Funding Statement

This study was funded by the Program Management Unit for Competitiveness Enhancement (PMU-C) National research, National Higher Education, Science, Research and Innovation Policy Council, Thailand through Clinixir Ltd.

### Author Declarations

The Central Research Ethics Committee (CREC) gave ethical approval for this work.

